# Epidemiology of CoVID-19 and predictors of recovery in the Republic of Korea

**DOI:** 10.1101/2020.05.07.20094094

**Authors:** Ashis Kumar Das, Saji Saraswathy Gopalan

## Abstract

**Background:** The recent CoVID-19 pandemic has emerged as a threat to global health. Though current evidence on the epidemiology of the disease is emerging, very little is known about the predictors of recovery.

**Objectives:** To describe the epidemiology of confirmed CoVID-19 patients in Republic of Korea and identify predictors of recovery.

**Materials and methods:** Using publicly available data for confirmed CoVID-19 cases from the Korea Centers for Disease Control and Prevention from January 20, 2020 to April 30, 2020, we undertook descriptive analyses of cases stratified by sex, age group, place of exposure, date of confirmation and province. Correlation was tested among all predictors (sex, age group, place of exposure and province) with the Pearson’s correlation coefficient. Associations between recovery from CoVID-19 and predictors were estimated using a multivariable logistic regression model.

**Results:** Majority of the confirmed cases were females (56%), from 20-29 age group (24.3%), and primarily from three provinces –– Gyeongsangbuk-do (36.9%), Gyeonggi-do (20.5%) and Seoul (17.1%). Case fatality ratio was 2.1% and 41.6% cases recovered. Older patients, patients from provinces such as Daegu, Gyeonggi-do, Gyeongsangbuk-do, Jeju-do, Jeollabuk-do and Jeollanam-do, and those contracting the disease from healthcare settings had lower recovery.

**Conclusions:** Our study adds to the very limited evidence base on potential predictors of survival among confirmed CoVID-19 cases. We call additional research to explore the predictors of recovery and support development of policies to protect the vulnerable patient groups.

## Introduction

For the first time, a novel coronavirus disease 2019 (CoVID-19) originating from Wuhan in China was reported to the World Health Organization in December of 2019.^1^ This novel coronavirus has taken the form of a major pandemic and has affected almost all major nations in the world. There have been more than 3.6 million confirmed cases and about 252,000 deaths as of May 05, 2020.^2^ The very first CoVID-19 case was diagnosed in the Republic of Korea (South Korea) on January 20, 2020.^3^ During the first two months of this global epidemic, South Korea had the second highest cases globally following China. According to the Korea Centers for Disease Control and Prevention (KCDC), there have been 10,804 confirmed cases and 254 deaths due to CoVID-19 as of May 5, 2020^4^

We present the epidemiology of CoVID-19 in the Republic of Korea using data from Korea Centers for Disease Control and Prevention and identify the predictors of recovery from the disease.

## Materials and methods

### Data source

The data were obtained from the Korea Centers for Disease Control and Prevention’s publicly shared sources. The dataset contains information about 3,388 confirmed COVID-19 cases in the Republic of Korea from January 20, 2020 through April 30, 2020. After excluding cases with missing values, 3,299 cases were included in the analysis.

### Variables

A confirmed case was defined as a person with laboratory confirmed positive test. The data contained the following patient details – age (in groups), sex, province, date of diagnosis, mode of exposure and outcome. There were 11 age groups – below 10 years, 10-19 years, 20-29 years, 30-39 years, 40-49 years, 50-59 years, 60-69 years, 70-79 years, 80-89 years, 90-99 years and above 100 years. We combined the last two age groups to create 90 years and above, and thus recategorized age to 10 groups. All seventeen provinces of the Republic of Korea were represented – Busan, Chungcheongbuk-do, Chungcheongnam-do, Daegu, Daejeon, Gangwon-do, Gwangju, Gyeonggi-do, Gyeongsangbuk-do, Gyeongsangnam-do, Incheon, Jeju-do, Jeollabuk-do, Jeollanam-do, Sejong, Seoul, and Ulsan. We categorized the dates of diagnosis by weeks, and they were as follows – 20-26 Jan 2020, 27 Jan-02 Feb 2020, 03-09 Feb 2020, 10-16 Feb 2020, 17-23 Feb 2020, 24 Feb-01 Mar 2020, 02-08 Mar 2020, 09-15 Mar 2020, 16-22 Mar 2020, 23-29 Mar 2020, 30 Mar-05 Apr 2020, 06-12 Apr 2020, 13-19 Apr 2020, 20-26 Apr 2020, 27-30 Apr 2020. Patients were exposed to potential CoVID-19 sources in multiple settings. The settings were nursing home, hospital, religious gathering, call center, community center, shelter and apartment, gym facility, overseas inflow, contact with patients and others. There were three outcomes – death, recovery and isolation. The confirmed patients after spending some days in isolation were retested. They were considered as recovered only after receiving a negative COVID-19 test.

### Statistical methods

We undertook descriptive analyses for the patient characteristics and presented the results stratified by sub-groups for each characteristic. Correlation was tested among all patient characteristics with the Pearson’s correlation coefficient. Associations between recovery from CoVID-19 and predictors (age group, sex, province and exposure) were estimated using a multivariable logistic regression model. We considered associations statistically significant if the p-value was below 0.05. The statistical analyses were performed using Python programming language Version 3.7 (Python Software Foundation, Wilmington, DE, USA) and Stata Version 15 (StataCorp LLC. College Station, TX).

## Results

### Pattern of the epidemic

As shown in figure 1, the first case of CoVID-19 was confirmed on January 20, 2020. There were a few daily cases of new infections for about a month. After a month, the curve suddenly rose starting February 19, 2020 to reach the peak around end of February and early March. It reached it peak on the 29^th^ of February with 813 confirmed cases. Though the curve descended after this date, still there were on an average 200 daily new confined cases until March 11, 2020. The curve continued its downward trend, however, adding at least 100 new daily cases through April 05, 2020. Towards the end of April, daily new confirmed cases were below 10.

**Figure 1.**
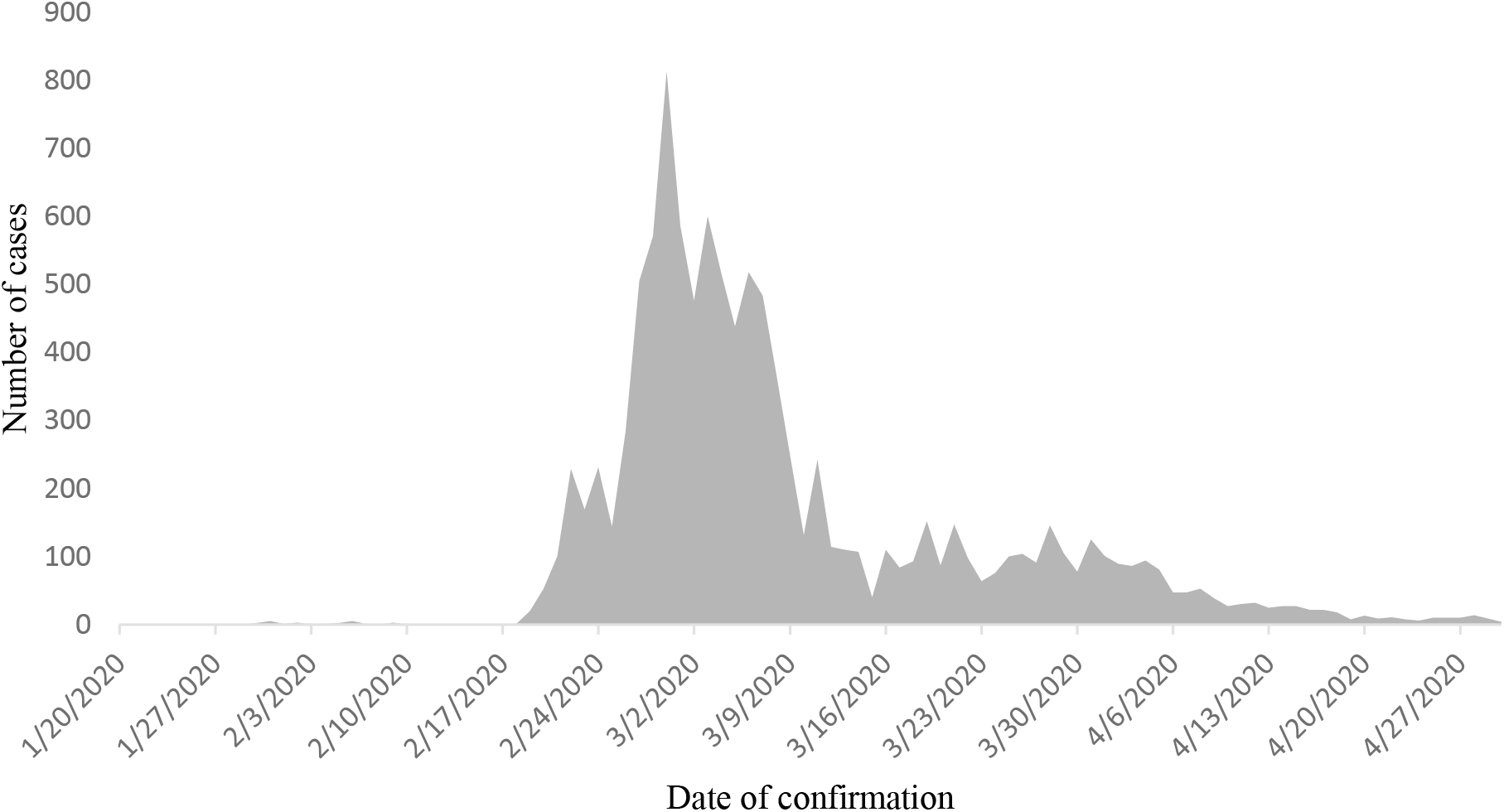
Daily new confirmed CoVID-19 cases in the Republic of Korea between January 20, 2020 and April 30, 2020

### Patient profile

Table 1 shows the profile of the patients. Out of 3,299 confirmed patients, a slightly more than half were females (56%). While there were cases from all age groups, the maximum patients were from 20-29 years (24.3%), followed by 50-59 years (18.1%), 40-49 years (13.8%), 30-39 years (13.3%) and 60-69 years (12.2%). Three provinces – Gyeongsangbuk-do (36.9%), Gyeonggi-do (20.5%) and Seoul (17.1%) – together accounted for the maximum patients. With respect to the exposure, it was unknown for the most (44%) followed by direct contact with patients (29%), from overseas (16.8%) and religious gathering (4.9%). According to this available data source, 85% percent of the patients were confirmed of their diagnosis between 24 February and 05 April of 2020. There were 61 deaths accounting for 2.1 percent (case fatality rate) of the patients. More than half were isolated (56.3%) and 41.6% recovered.

**Table 1.**
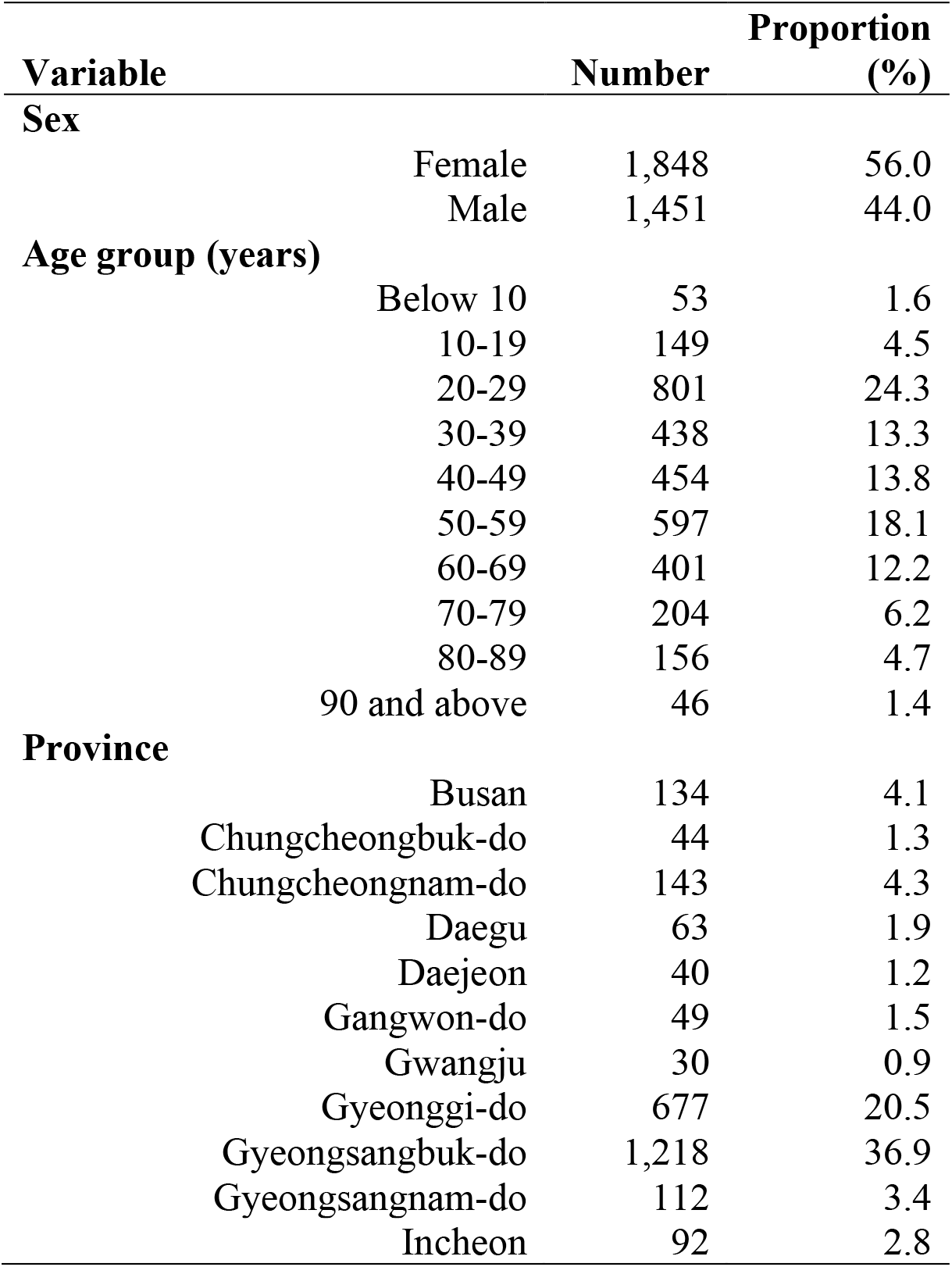
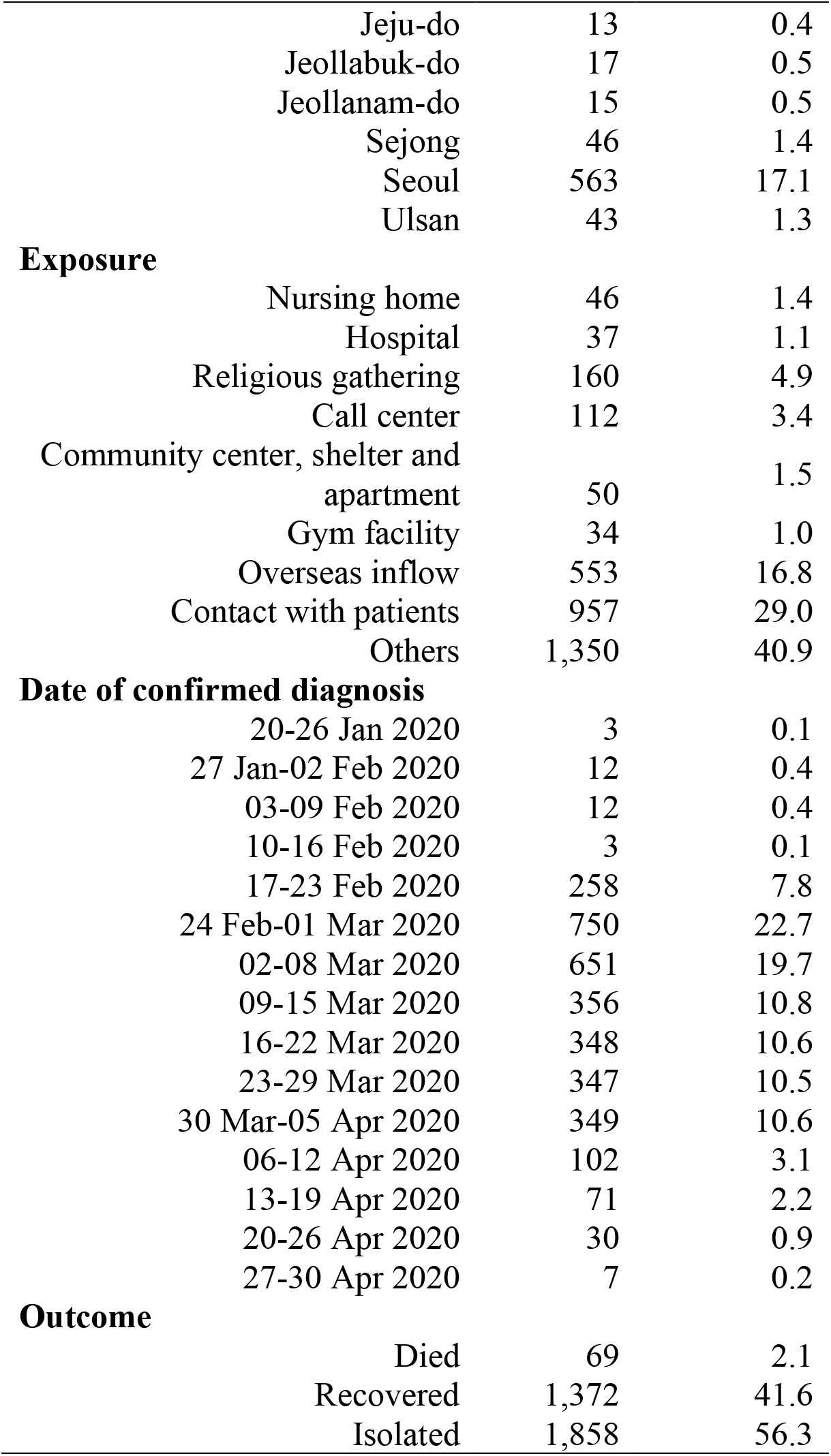
Sample characteristics (N=3,299)

| **Variable** | **Number** | **Proportion (%)** |
| --- | --- | --- |
| **Sex** |  |  |
| Female | 1,848 | 56.0 |
| Male | 1,451 | 44.0 |
| **Age group (years)** |  |  |
| Below 10 | 53 | 1.6 |
| 10-19 | 149 | 4.5 |
| 20-29 | 801 | 24.3 |
| 30-39 | 438 | 13.3 |
| 40-49 | 454 | 13.8 |
| 50-59 | 597 | 18.1 |
| 60-69 | 401 | 12.2 |
| 70-79 | 204 | 6.2 |
| 80-89 | 156 | 4.7 |
| 90 and above | 46 | 1.4 |
| **Province** |  |  |
| Busan | 134 | 4.1 |
| Chungcheongbuk-do | 44 | 1.3 |
| Chungcheongnam-do | 143 | 4.3 |
| Daegu | 63 | 1.9 |
| Daejeon | 40 | 1.2 |
| Gangwon-do | 49 | 1.5 |
| Gwangju | 30 | 0.9 |
| Gyeonggi-do | 677 | 20.5 |
| Gyeongsangbuk-do | 1,218 | 36.9 |
| Gyeongsangnam-do | 112 | 3.4 |
| Incheon | 92 | 2.8 |

|  |  |  |  |
| --- | --- | --- | --- |
|  | Jeju-do | 13 | 0.4 |
|  | Jeollabuk-do | 17 | 0.5 |
|  | Jeollanam-do | 15 | 0.5 |
|  | Sejong | 46 | 1.4 |
|  | Seoul | 563 | 17.1 |
|  | Ulsan | 43 | 1.3 |
| **Exposure** |  |  |  |
|  | Nursing home | 46 | 1.4 |
|  | Hospital | 37 | 1.1 |
|  | Religious gathering | 160 | 4.9 |
|  | Call center | 112 | 3.4 |
|  | Community center, shelter and apartment | 50 | 1.5 |
|  | Gym facility | 34 | 1.0 |
|  | Overseas inflow | 553 | 16.8 |
|  | Contact with patients | 957 | 29.0 |
|  | Others | 1,350 | 40.9 |
| **Date of confirmed diagnosis** |  |  |  |
|  | 20-26 Jan 2020 | 3 | 0.1 |
|  | 27 Jan-02 Feb 2020 | 12 | 0.4 |
|  | 03-09 Feb 2020 | 12 | 0.4 |
|  | 10-16 Feb 2020 | 3 | 0.1 |
|  | 17-23 Feb 2020 | 258 | 7.8 |
|  | 24 Feb-01 Mar 2020 | 750 | 22.7 |
|  | 02-08 Mar 2020 | 651 | 19.7 |
|  | 09-15 Mar 2020 | 356 | 10.8 |
|  | 16-22 Mar 2020 | 348 | 10.6 |
|  | 23-29 Mar 2020 | 347 | 10.5 |
|  | 30 Mar-05 Apr 2020 | 349 | 10.6 |
|  | 06-12 Apr 2020 | 102 | 3.1 |
|  | 13-19 Apr 2020 | 71 | 2.2 |
|  | 20-26 Apr 2020 | 30 | 0.9 |
|  | 27-30 Apr 2020 | 7 | 0.2 |
| **Outcome** |  |  |  |
|  | Died | 69 | 2.1 |
|  | Recovered | 1,372 | 41.6 |
|  | Isolated | 1,858 | 56.3 |

### Predictors of recovery

As shown in figure 2, there were strong no correlations between the predictors. Compared to younger age groups (table 2), older patients had lower recovery – 70-79 years (adjusted odds ratio 0.31; p value 0.01), 80-89 years (aOR 0.22; p value 0.001) and 90 years and above (aOR 0.13; p value <0.001). Provinces such as Daegu, Gyeonggi-do, Gyeongsangbuk-do, Jeju-do, Jeollabuk-do and Jeollanam-do had statistically significant lower recovery rates than Busan. When compared with exposure from nursing homes, patients who were exposed to COVID-19 infection from religious gatherings, community dwellings, and others had higher recovery rates.

**Figure 2.**
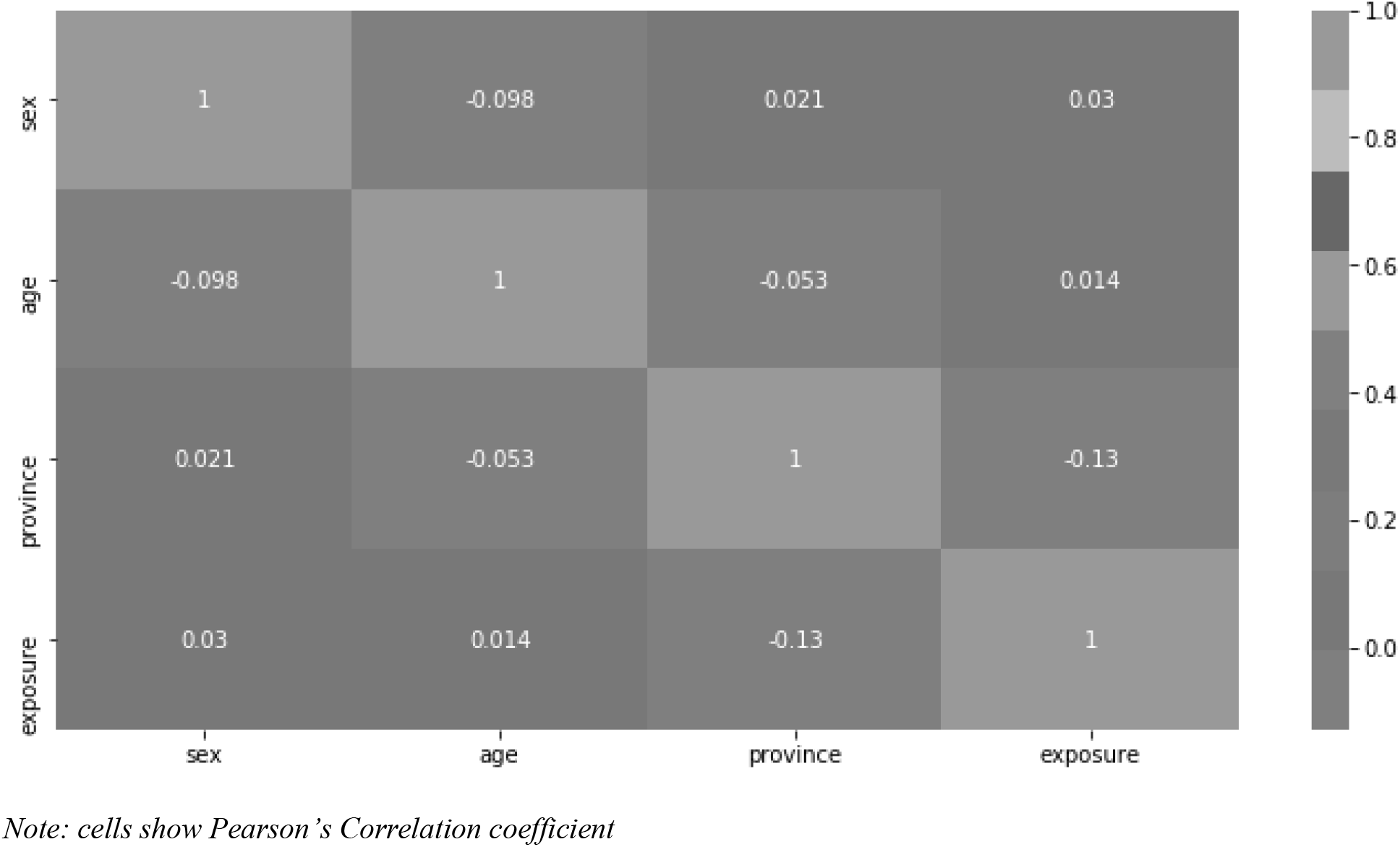
Correlation among predictors

**Table 2.** Predictors of recovery

| Variable | Recovery (%) | Odds ratio | 95% confidence interval | p value |
| --- | --- | --- | --- | --- |
| **Sex** |  |  |  |  |
| Female | 56.6 | Reference |  |  |
| Male | 56.0 | 0.90 | 0.73-1.10 | 0.312 |
| **Age group (years)** |  |  |  |  |
| Below 10 | 60.4 | Reference |  |  |
| 10-19 | 58.4 | 1.33 | 0.55-3.25 | 0.527 |
| 20-29 | 46.8 | 1.51 | 0.68-3.36 | 0.314 |
| 30-39 | 50.7 | 1.77 | 0.78-4.04 | 0.173 |
| 40-49 | 38.8 | 1.13 | 0.50-2.56 | 0.771 |
| 50-59 | 39.5 | 1.02 | 0.45-2.31 | 0.955 |
| 60-69 | 44.4 | 0.81 | 0.35-1.84 | 0.611 |
| 70-79 | 38.7 | 0.31 | 0.13-0.76 | 0.01 |
| 80-89 | 32.7 | 0.22 | 0.09-0.54 | 0.001 |
| 90 and above | 32.6 | 0.13 | 0.04-0.37 | <0.001 |
| **Province** |  |  |  |  |
| Busan | 82.8 | Reference |  |  |
| Chungcheongbuk-do | 88.6 | 1.02 | 0.35-2.99 | 0.971 |
| Chungcheongnam-do | 88.8 | 1.38 | 0.61-3.12 | 0.432 |
| Daegu | 6.4 | 0.00 | 0.00-0.01 | <0.001 |
| Daejeon | 85.0 | 1.38 | 0.43-4.44 | 0.585 |
| Gangwon-do | 59.2 | 0.30 | 0.12-0.78 | 0.013 |
| Gwangju | 70.0 | 0.67 | 0.25-1.79 | 0.424 |
| Gyeonggi-do | 9.0 | 0.01 | 0.01-0.02 | <0.001 |
| Gyeongsangbuk-do | 63.5 | 0.10 | 0.05-0.19 | <0.001 |
| Gyeongsangnam-do | 85.7 | 0.93 | 0.40-2.15 | 0.866 |
| Incheon | 59.8 | 0.55 | 0.26-1.18 | 0.125 |
| Jeju-do | 53.9 | 0.29 | 0.08-1.00 | 0.05 |
| Jeollabuk-do | 23.5 | 0.03 | 0.01-0.11 | <0.001 |
| Jeollanam-do | 20.0 | 0.03 | 0.01-0.13 | <0.001 |
| Sejong | 87.0 | 1.20 | 0.25-5.86 | 0.821 |
| Seoul | 74.1 | 0.70 | 0.38-1.29 | 0.257 |
| Ulsan | 86.1 | 1.24 | 0.32-4.83 | 0.76 |
| **Exposure** |  |  |  |  |
| Nursing home | 17.4 | Reference |  |  |
| Hospital | 43.2 | 1.14 | 0.34-3.85 | 0.833 |
| Religious gathering | 80.6 | 6.29 | 1.80-21.94 | 0.004 |
| Call center | 84.8 | 2.68 | 0.77-9.31 | 0.122 |
| Community center, shelter and apartment | 82.0 | 13.34 | 3.06-58.05 | 0.001 |
| Gym facility | 94.1 | 6.05 | 0.63-57.79 | 0.118 |
| Overseas inflow | 40.9 | 2.71 | 0.90-8.13 | 0.075 |
| Contact with patients | 41.5 | 2.80 | 0.98-8.03 | 0.055 |
| Others | 67.7 | 7.14 | 2.58-19.75 | <0.001 |

## Discussion

Due to multipronged approaches (proactive surveillance, higher testing, isolation, quarantine, use of technology, masks and social distancing campaigns) by the government, incidence of new cases came down sharply in South Korea by mid-March and further to less than 10 new cases by mid-April.^5^

Our study shows that females constituted the majority of confirmed cases, whereas males accounted for most of the confirmed cases in China and Italy.^6–9^ Around a fourth of the cases were from 20-29 years age group unlike in most other countries where the infected were older.^6,7^ The possible reason for higher representation of younger population in our sample could be specific exposure to cluster of cases through participation in religious activities or workplaces.^5,10^ The case fatality rate was much lower (2.1%) compared to other countries such as Italy (13.3%) and China (4%). Similar to findings from several other countries, we found the elderly to be more vulnerable with lower probabilities of recovery.^6,8,11>^ It is quite possible that presence of pre-existing medical conditions in elderly predispose them to delayed recovery. We also found cases contracting the infection in non-healthcare setting had higher recovery. While there is no such evidence currently, there could be a possibility that the exposure outside non-healthcare setting might have involved relative younger and healthier cases. Considering our study findings, we suggest additional measures to protect the vulnerable cases who are less likely to recover from the infection. Thus, elderly and cases contracting infection from healthcare settings should be given special attention.

Our study has two potential limitations. First, we used publicly available data of only a third of confirmed cases in the country. Thus, we are unable to ascertain the representativeness of the data for all confirmed cases in South Korea. So, the findings will have to be interpreted with caution. Secondly, the data lacks information of patients’ symptoms and clinical features. Inclusion of these potential predictors would have enhanced the relevance of this study further. Despite these limitations, our study adds to the very limited evidence base on potential predictors of recovery among confirmed CoVID-19 cases.^12^ However, we believe the evidence base be strengthened with further relevant research as authorities make more data publicly available or through primary hospital based studies.

## Conclusions

The CoVID-19 pandemic has emerged as a great threat to global health challenging health systems across the world to efficiently deal with this situation. Emerging evidence on vulnerability to COVID-19 and predictors of recovery will inform providers and policy makers to effectively triage and prioritize limited resources. Therefore, we call for additional research to explore the predictors of recovery and support development of policies to protect the vulnerable patient groups.

## Data Availability

Study uses publicly available data from Korea CDC

## Data Availability

The data used to support the findings of this study are available publicly through the Korea Centers for Disease Control and Prevention.

## Acknowledgements

We are grateful to Korea Centers for Disease Control and Prevention for making this data publicly available.

